# Are Electromyography data a fingerprint for patients with cerebral palsy (CP)?

**DOI:** 10.1101/2024.09.22.24314168

**Authors:** Mehrdad Davoudi, Firooz Salami, Robert Reisig, Dimitrios A. Patikas, Nicholas A. Beckmann, Katharina Susanne Gather, Sebastian I. Wolf

**Author notes:** Correspondence: Sebastian I. Wolf Clinic for Orthopedics, Schlierbacher Landstr. 200a, 69118 Heidelberg, Germany.

## Abstract

This study aimed to first investigate changes in electromyography (EMG) patterns after multilevel surgical treatment in patients with cerebral palsy (CP) and then to assess the connection between the measure of EMG and motor control indices and surgery outcomes. We analyzed retrospective EMG and gait data from 167 patients with CP before and after surgery and from 117 typically developed individuals as a reference group. The patients underwent at least one soft tissue surgery on their shank and foot muscles. Using Repeated Measures ANOVA, we examined the norm-distance (ND) of the kinematics, kinetics, and EMG patterns, in addition to the Kerpape-Rennes EMG-based Gait Index (EDI), EMG Profile Score (EPS), and Walking Dynamic Motor Control Index (DMC) before and after surgery. Participants were divided into different response groups (Poor, Mild, and Good gait quality) according to their pre- and post-treatment Gait Deviation Index (GDI), using the K-means-PSO clustering algorithm. The gait and EMG indices were compared between the responders using the nonparametric Mann-Whitney test. The ND for all kinematics and kinetics parameters significantly improved (p-value < 0.05) after the surgery. Regarding EMG, a significant reduction was only observed in the ND of the rectus femoris (p-value < 0.001) and soleus (p-value = 0.006). Among the indices, DMC was not altered post-operatively (p-value = 0.88). Although EDI and EPS were consistent across responders with a similar pre-treatment gait, a higher DMC was significantly associated with a greater improvement, particularly in patients with poor gait (p-value < 0.05). These findings indicate systematic changes in the EMG of patients with CP following surgery, which can also be demonstrated through indices. DMC is a measure that can potentially serve as a partial predictor of outcomes, particularly in patients with poor pre-operative gait. Future research should investigate the effects of different surgical strategies on the improvement of these patients.

## 1. Introduction

Does the electromyogram (EMG) remain unchanged as a fingerprint in patients with cerebral palsy (CP) after orthopedic surgery? Visually inspecting pre- and post-operative EMG signals in patients with CP, Gueth et al. [1] answered this question as a ‘yes’, indicating that EMG is robust to surgery. Therefore, they proposed the idea that we don’t need to measure the EMG after the operation. However, the assessment of a small number of patients as well as subjectivity are two main limitations of their study.

Patikas et al. delved deeper into this question by monitoring 34 patients, focusing on the changes in envelopes of EMG data rather than raw signals [2]. They observed a systematic improvement in the EMG of shank muscles (soleus, tibialis anterior, and lateral gastrocnemius) after a single-event multilevel surgery (SEMLS). However, the reported changes compared to kinematics and kinetics were relatively small. The majority of the patients in their study had equinus foot deformity with a history of calf muscle lengthening surgery, indicating the significant impact of shank soft tissue surgery on EMG in patients with CP. These findings aligned with other studies that indicated muscle activation patterns, as well as their recruitment in patients with CP, may change after an intervention [3–5]. Consequently, they concluded that EMG can describe the clinical condition of the patient before and after orthopedic surgery, and it can be considered for better clinical decision-making, such as the approach developed by Reinbold et al. [6] to predict the results of rectus transfer surgery.

The suggestion from Patikas et al. [2], which served as the main motivation for the current study, was to conduct a controlled study with a larger population and more homogeneous surgical treatment. They argued that employing such standardized approaches could help clarify the pre-operative compensatory mechanisms present in patients, ultimately improving the prediction of surgical intervention outcomes. Nonetheless, there remains a need for the development of new analytical methods to interpret EMG signals effectively in a clinical context. Moreover, the subjective evaluation of the pre-operative motor control status’ impact on surgical outcomes, in conjunction with CP biomechanics, constitutes a notable gap in extant literature warranting attention.

Using clustering analysis, Davoudi et al. [7, 8] established an association between the pre-operation activity of muscles in patients with CP and their response to the surgery. Moreover, they introduced a simple index as the ratio between the activity of the rectus femoris and gastrocnemius lateralis muscles as a predictor of the chance of improvement in patients with a crouch gait. Kinematic indices such as the Gait Deviation Index (GDI) [9] and the Gait Profile Score (GPS) [10] are two measures of gait quality whose applicability in the assessment of gait with CP has already been reported in the literature [11–14]. The Kerpape-Rennes EMG-based Gait Index [15], referred to as the EMG Deviation Index (EDI) in this study, and the EMG Profile Score (EPS) [16] are two EMG-based measures calculated using the same methodology as the GDI and GPS, respectively. EDI quantifies global muscle activity based on the Euclidean distance between a patient’s EMG pattern and that of typically developing (TD) subjects using principal component analysis. Scores equal to or above 100 indicate normal gait, with a decrease of 10 points indicating a deviation of one standard deviation (SD) from TD. Additionally, to assess deviations from the norm for individual muscle groups independently, the EPS was calculated based on the mean of the root mean square error (RMSE) for each muscle during the gait, providing a score without units. While these are validated measures [17], there is no study on the use of EMG indices in the CP population.

Although EDI and EPS can describe biomechanics by considering patterns, they are relatively weak in presenting the degree of motor involvement of the patient. To address this limitation, Schwartz et al. introduced the Walking Dynamic Motor Control Index (Walk-DMC) to clinically assess the effect of altered neuromuscular control on treatment outcomes in patients with CP [18]. A higher DMC suggests better motor function, with 100 being the average DMC for TD individuals, and 10 points representing one standard deviation from typical development outcomes. The observed DMC was significantly associated with the response to treatment, second only to the pre-treatment level of GDI. However, the treatment in their study included various interventions such as surgery, physical therapy, and selective dorsal rhizotomy. Additionally, the influence of DMC level on individuals with similar gait quality prior to treatment was not investigated. Therefore, it remains unclear how DMC can be helpful for predicting the results of surgery in patients with CP.

The primary aim of this study was to investigate the effect of SEMLS on the EMG and gait kinematics of patients with CP. To achieve a more homogeneous surgical management across the cohort, we focused on patients who underwent soft tissue surgery on their shank and foot muscles, in addition to potentially other more proximal surgeries. Furthermore, we evaluated the applicability of EMG indices as global measures for assessing the results of surgery in the CP population. To the best of our knowledge, this is the first time EDI and EPS have been measured in patients with CP. Moreover, considering DMC, we assessed the connection between motor control and surgery outcomes for subgroups with the same level of GDI. We hypothesized that while EMG patterns, as a biomechanical aspect of EMG activity, will change with surgery, motor control is rather independent of a specific gait pattern and remains stable. Our second hypothesis is that DMC is robust against biomechanical correction following surgery and potentially, this measure before treatment can serve as a predictor for the level of improvement after SEMLS in patients with CP.

## 2. Methods

### 2.1. Ethics statement

The study was approved by the local Ethical Committee “Medical Faculty, Heidelberg University (no: S-243/2022)”.

### 2.2. Participants

The data analyzed in this retrospective study were part of a larger database established at the local University Clinics in the years 2000-2022 when retrieval was stopped. Only personnel that had regular legal access to the medical records retrieved patient data, collected and anonymized it. After this step, individual participants could not be identified anymore. They collected data in the time November and December 2022, and anonymized it in the same year December 28^th^.

The database was filtered for CP patients with at least two consecutive examinations in the local gait laboratory with orthopedic surgery in between. Examinations that did not show high-quality EMG data were excluded, and surgeries had to address gait disorders caused by CP. If multiple examinations were available, the dates closest to the surgery were selected. The first examination (E1) was conducted before the operation, and the second examination (E2) was typically conducted one year after the operation. Therefore, E1 and E2 pertain to the same individuals evaluated at different time points. For hemiplegia patients, only data from the affected side were considered.

Further primary inclusion criteria encompassed gait and clinical data for each examination, walking barefoot without assistive devices, and classification as Gross Motor Function Classification System (GMFCS) level I or II. Usually, the surgeries involved multiple procedures on different levels (hip, thigh, shank, and feet). If multiple surgeries were performed on different dates between the two examinations, all procedures were included in this study as they could be relevant for the second examination. Following the recommendation of Patikas et al. [2], to ensure a more homogeneous approach to surgical treatment, we focused on patients who, among other procedures, underwent soft tissue surgery on muscles located at the shank and foot level. For 91% of patients, this involved at least one surgery on the triceps surae muscle, such as the Baumann Procedure [19] (46%), Strayer Procedure [20] (46%), or Achilles Tendon Lengthening [21] (17%). Additionally, there were some muscle transfer surgeries, such as tibialis anterior transfer (13%) and tibialis posterior transfer (9%), and multiple muscle lengthening procedures, such as flexor digitorum longus lengthening (3%). In most cases, the surgery included further procedures on another level, i.e., legs and feet. The most prevalent additional procedures were femoral derotation (78%), bony foot procedures (38%), rectus transfer (30%), and hamstring lengthening (28%).

Furthermore, according to [2], we chose to assess only the more involved side in each patient to maintain homogeneity regarding severity. This was defined as the side that underwent surgery, and in cases of bilateral involvement, the side with the lower GDI was selected for further analysis. Following the application of these criteria, 167 patients were recruited, along with 117 TD individuals serving as the reference group. Table 1 presents the characteristics of the study participants.

**Table 1.** Demographic and descriptive data of the participants at their first (E1) and second (E2) examinations and also TD individuals.

|  | **E1 (n=167)**<br>**pre-operative examination** | **E2 (n=167)**<br>**post-operative examination** | **TD (n=117)** |
| --- | --- | --- | --- |
| Age (years) | 15.86± 8.6 (5.3-49.5) | 18.1 ± 8.9 (6.9-54.2) | 21.7 ± 12.3 (6.0-46.0) |
| Height (cm) | 149.1 ± 18.5 (105.0-194.0) | 152.6 ± 18.2 (121.0-187) | 161.9 ± 19.8 (108.0-195.0) |
| Body mass (kg) | 43.68 ± 16 (14.6-87.3) | 46.8 ± 17.3 (14.8-101.8) | 66.4 ± 16.8 (19.0-91.0) |
| Sex (male/female) | 84/83 |  | 58/59 |
| CP type (diplegia/ hemiplegia) | 148/19 |  |  |
| Interval between examinations (years) | 2.3 ± 1.8 (0.8-13.5) |  |  |

### 2.3. Data processing

The data recording and processing approach used in this study was the same as the one described in our recently published paper on rectus femoris EMG clustering [7]. Envelopes [22] for the EMG of seven major lower-extremity muscles—rectus femoris, vastus lateralis, tibialis anterior, semimembranosus, biceps femoris, lateral gastrocnemius, and soleus—were extracted for subsequent analysis. Additionally, we calculated the kinematics, kinetics, and spatiotemporal parameters of participants’ gait.

To quantify the extent of deviation in a patient’s EMG envelopes and gait parameters relative to a reference group, we calculated the norm-distance (ND) according to [2]. ND was defined as the absolute difference between a muscle’s EMG envelope at the i_th data point belongs to the patient p (*F*_*pi*_) and the mean value of the corresponding data point within that muscle for the reference group (*F̅*_*ni*_), divided by the respective standard deviation within the reference group (*SD*_*ni*_), as expressed in Equation (1). The ND values for all 101 data points were averaged over a gait cycle for further analysis. Changes in the average ND (*N̅D̅*) from the initial examination (E1) to the subsequent examination (E2) were regarded as indicative of the surgery’s impact on the patient’s EMG. A reduction in *N̅D̅* following the intervention suggested an improvement towards a pattern more similar to the reference group.

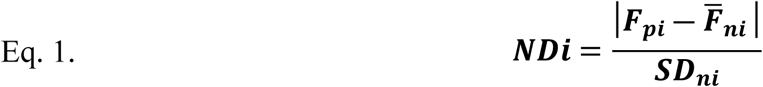

The same procedure was also employed to assess gait patterns in sagittal plane, including angles, moments, and power at the hip, knee, and ankle joints.

### 2.4. Gait and EMG Indices

EMG indices, including EDI [15], EPS [16], and Walk-DMC [18], were derived from the envelopes in accordance with existing literature. An increase in EDI, along with a reduction in EPS, may signify improvement post-surgery. Moreover, a higher DMC indicates enhanced motor control function in the patient. Additionally, GDI [9] and GPS [10], as measures of gait quality, were evaluated in both E1 and E2 for the patients. A higher GDI and a lower GPS are associated with reduced deviation from a typical gait pattern. All analyses for the extraction of parameters and indices were conducted using Matlab software (The MathWorks, Inc., USA).

### 2.5. Clustering

We utilized the same k-means-PSO clustering algorithm as in our previous study [7] to categorize patients into three different gait levels based on their GDI in each examination, pre-(E1) and post-operative (E2). By averaging the GDI of patients within each cluster, we classified the patients as having good, mild, and poor gait quality, corresponding to high, medium, and low mean GDI, respectively (Table 4 in the results section). Figure 1 shows the clustering procedure applied to our database. The clustering algorithm was developed using Matlab, based on the details described in [7]. The algorithm was identical for both E1 and E2 populations. The number of clusters was determined by us in a supervised manner, set at ’n=3’. This allowed us to track the changes in gait quality of the patients (Figure 1). Although the patients in both E1 and E2 are the same (matched), their GDI values may differ due to the effects of the surgery (Figure 1.A and B).

**Fig 1.**
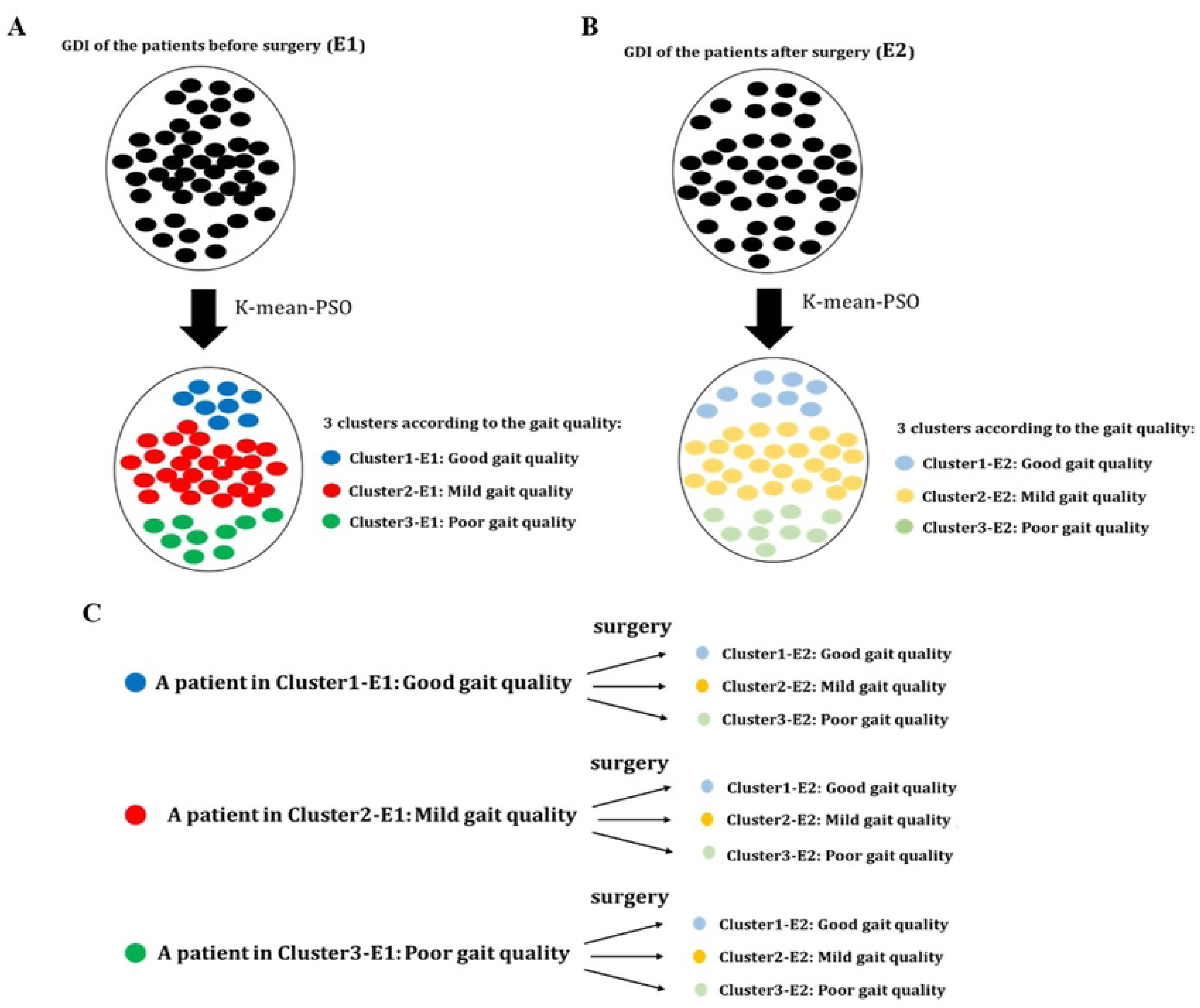
The clustering of the patients, (A) according to their pre-operation (E1) GDI, and (B) according to their post-operation (E2) GDI. (C) The possible responses of the patients to the surgery, identified within each cluster both pre- and post-operation.

Using this approach, we removed the effect of the pre-treatment level of GDI, which, according to [18], significantly influences the outcomes of surgery. Subsequently, we identified three groups of patients with the same pre-operative GDI level (E1). Each patient could have one of three possible responses to the intervention: good, mild, or poor, corresponding to being identified as Cluster 1, Cluster 2, or Cluster 3 post-operatively (E2) (Figure 1.C). Furthermore, the specifics of the type and number of surgeries performed between E1 and E2 for different responders are examined to investigate any potential bias arising from the treatment approach on the responses and clustering outcomes. According to the surgical details described in section 2.2, the most frequent proximal surgeries (femoral derotation, rectus transfer, and hamstring lengthening) and the main distal surgeries (Baumann and Strayer procedures, Achilles tendon lengthening, and bony foot procedures) were considered for examination between the clusters.

### 2.6. Statistics

To compare the effect of the intervention on the ND of kinematics, kinetics, and EMG parameters (sections 2-3) and on the indices outlined in section 2-4, Repeated Measures ANOVA was used. Further, for the muscles that showed a significant improvement (p-value = 0.05) in their ND, we applied statistical parametric mapping (SPM, www.spm1d.org) implemented in Matlab [23] to compare the changes over the entire gait cycle. We applied the nonparametric Mann-Whitney test (p-value = 0.05) to compare the primary levels of the gait and EMG indices between the groups with the same condition in E1, and also to assess their changes from E1 to E2.

## 3. Results

The ND of all measured kinematics and kinetics parameters exhibited significant improvement (p-value < 0.05) post-surgery, as shown in Table 2. Regarding EMG, a significant reduction was only observed in the ND of the EMG for the rectus femoris (p-value < 0.001) and soleus (p-value = 0.006), as detailed in Table 3. While the gait indices (GDI and GPS) and EMG indices (EDI and EPS) demonstrated significant changes towards normal values from E1 to E2 (p-value < 0.001), the measure for motor control (DMC) showed no significant difference after the intervention (p-value = 0.88).

**Table 2.** Pre- (E1) and post-operative (E2) norm-distance of the kinematics and kinetics parameters and for GDI and GPS.

| Measure | Examination | Mean (SD) | 95% Confidence interval<br>(lower bound - upper bound) | Sig. |
| --- | --- | --- | --- | --- |
| ND ankle dorsiflexion (degree) | E1 | 3.36 (0.22) | 2.93 - 3.78 | <0.001 |
|  | E2 | 1.81 (0.07) | 1.67 - 1.95 |  |
| ND ankle dorsiflexion moment (Nm/kg) | E1 | 2.07 (0.04) | 1.99 - 2.15 | <0.001 |
|  | E2 | 1.63 (0.04) | 1.54 - 1.71 |  |
| ND ankle dorsiflexion power (W/kg) | E1 | 1.80 (0.06) | 1.68 - 1.91 | <0.001 |
|  | E2 | 1.29 (0.04) | 1.21 - 1.38 |  |
| ND knee flexion (degree) | E1 | 3.24 (0.14) | 2.96 - 3.52 | <0.001 |
|  | E2 | 2.53 (0.12) | 2.28 - 2.77 |  |
| ND knee flexion moment (Nm/kg) | E1 | 1.76 (0.06) | 1.65 - 1.87 | <0.001 |
|  | E2 | 1.55 (0.05) | 1.44 - 1.66 |  |
| ND knee flexion power (W/kg) | E1 | 1.24 (0.03) | 1.18 - 1.30 | <0.001 |
|  | E2 | 1.11 (0.02) | 1.07 - 1.16 |  |
| ND hip flexion angle (degree) | E1 | 1.81 (0.08) | 1.66 - 1.97 | <0.001 |
|  | E2 | 1.52 (0.07) | 1.39 - 1.65 |  |
| ND hip flexion moment (Nm/kg) | E1 | 1.85 (0.05) | 1.76 - 1.95 | <0.001 |
|  | E2 | 1.54 (0.05) | 1.45 - 1.64 |  |
| ND hip flexion power (W/kg) | E1 | 1.74 (0.04) | 1.67 - 1.81 | <0.001 |
|  | E2 | 1.55 (0.04) | 1.47 - 1.63 |  |
| GDI | E1 | 58.51 (0.96) | 56.61 - 60.42 | <0.001 |
|  | E2 | 73.74 (0.92) | 71.93 - 75.55 |  |
| GPS | E1 | 14.35 (0.35) | 13.66 - 15.03 | <0.001 |
|  | E2 | 9.73 (0.24) | 9.27 - 10.20 |  |

**Table 3.**
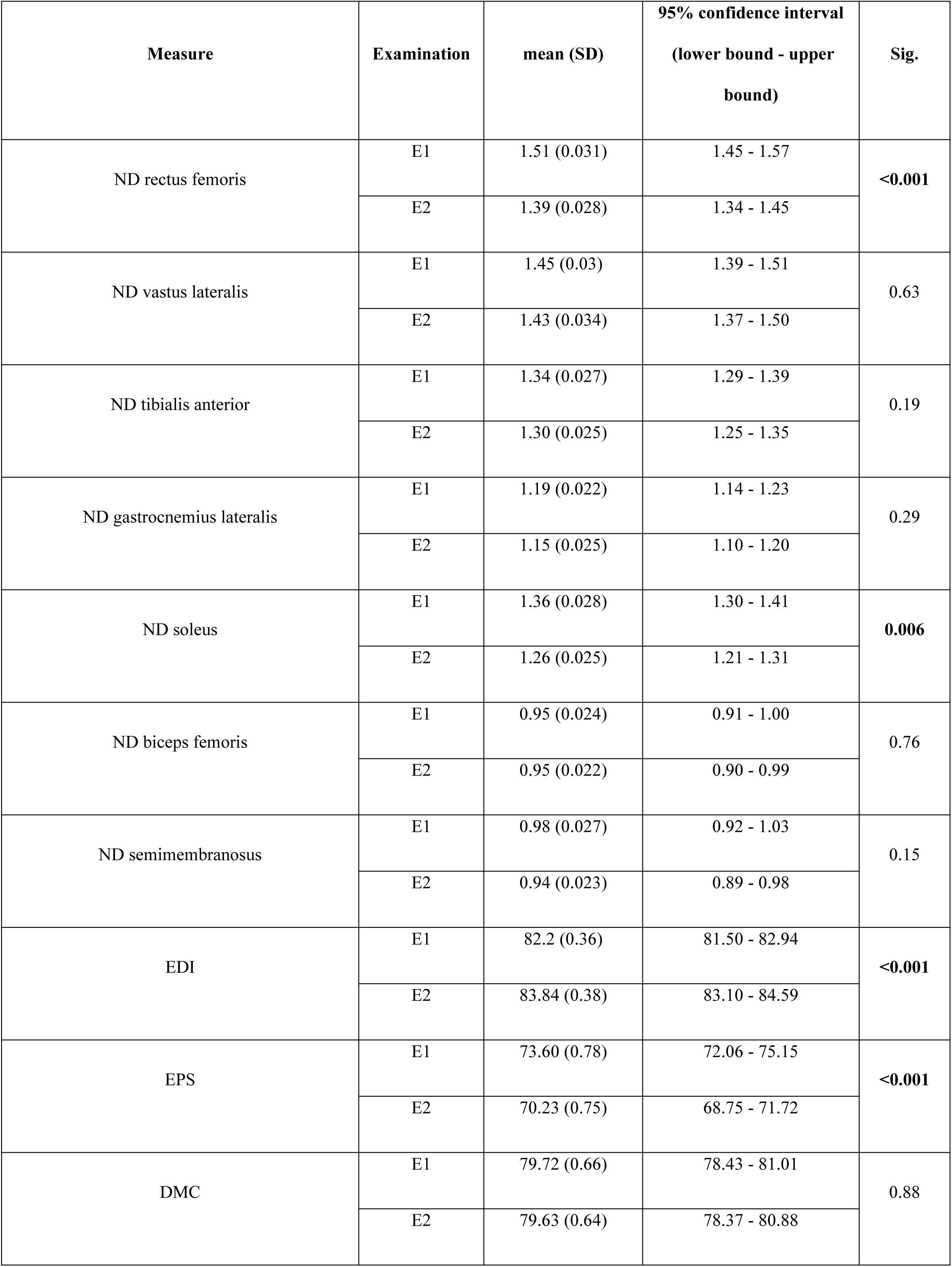
Pre- (E1) and post-operative (E2) norm-distance of the EMG activity of muscles and for EDI, EPS and DMC.

**Table 4.** Average GDI of the patients within each identified cluster pre- and post-operation, along with the assigned gait conditions.

| Examination time | Cluster | Average (SD) of GDI for the patients in the cluster | Assigned gait condition |
| --- | --- | --- | --- |
| **E1 (pre-operation)** | Cluster1 | 71.02 (6.9) | Good |
|  | Cluster2 | 55.18 (3.0) | Mild |
|  | Cluster3 | 43.43 (5.0) | Poor |
| **E2 (post-operation)** | Cluster1 | 86.47 (6.8) | Good |
|  | Cluster2 | 72.28 (3.3) | Mild |
|  | Cluster3 | 58.51 (6.7) | Poor |

SPM analysis revealed significant differences at specific phases of the gait cycle for the rectus femoris, approximately 20% (early stance) and 75% (mid-swing) (Figure 2.A). For the soleus, significant differences were observed at the beginning (initial contact), around 50% (end of stance), and at the end (terminal swing) of the gait cycle between E1 and E2 (Figure 2.B).

**Fig. 2.**
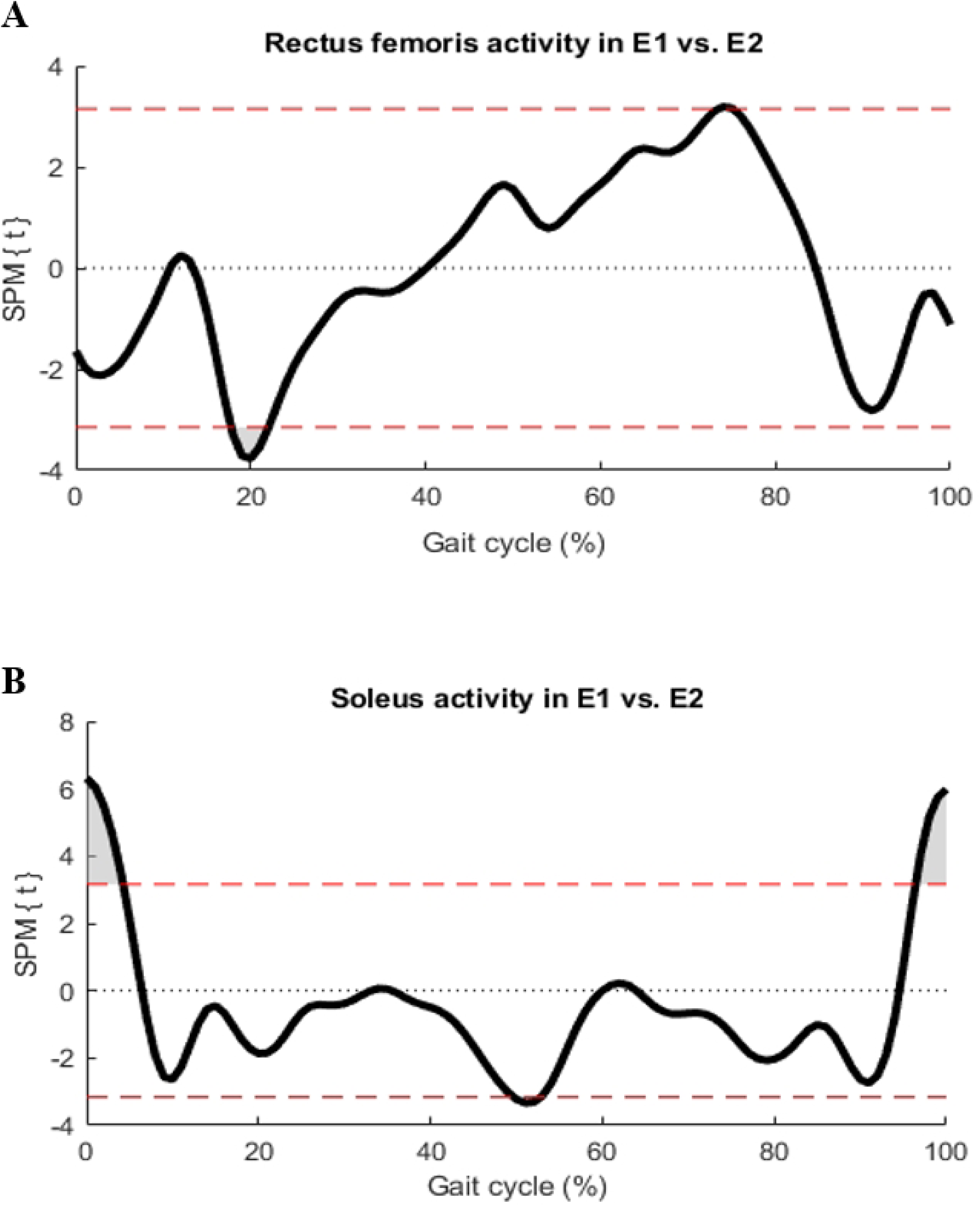
Results of SPM analysis highlighting differences in EMG patterns of rectus femoris (A) and soleus (B) muscles pre-operation (E1) and post-operation (E2) throughout the gait cycle. Dark areas indicate significant differences in activity.

Initial clustering results provided in Table 4 indicate the mean and SD of the GDI for patients identified in each cluster at two assessment points (pre- and post-operation). Patients with the best and the poorest gait performance at E1 had an average GDI of 71.02 and 43.43, respectively. The same assigned gait conditions had an average GDI of 86.47 and 58.51 at E2. This illustrates the general effect of the surgery on improving gait, as evidenced by the increase in the mean GDI from 58.51 at E1 to 73.74 at E2. However, since individual responses varied, patients were categorized according to the change in their cluster post-operation. Tables 5, 6, and 7 compare those with similar pre-operative gait, minimizing the influence of initial gait quality.

**Table 5.** Comparative analysis of gait and EMG indices among participants with poor gait quality at baseline (E1) across responders according to their post-operation gait quality.

| Variable | Gait quality condition before surgery (E1) | Gait quality condition after surgery (E2) | No. | Mean (SD) in E1 | Mean (SD) in E2 | P-Value between E1-E2 | P-Value between Poor_to_Good and Poor_to_Mild in E1 | P-Value between Poor_to_Good and Poor_to_Poor in E1 | P-Value between Poor_to_Mild and Poor_to_Poor in E1 |
| --- | --- | --- | --- | --- | --- | --- | --- | --- | --- |
| GDI | Poor | Good | 10 | 42.27 (5.8) | 86.21 (7.4) | <0.001 | 0.418 | 0.387 | 0.516 |
|  | Poor | Mild | 22 | 43.98 (4.8) | 72.55 (0.8) | <0.001 |  |  |  |
|  | Poor | Poor | 11 | 43.56 (5.2) | 57.01 (8.0) | <0.001 |  |  |  |
| GPS | Poor | Good | 10 | 20.92 (3.4) | 6.93 (1.2) | <0.001 | 0.392 | 0.453 | 0.422 |
|  | Poor | Mild | 22 | 19.96 (2.7) | 9.54 (0.98) | <0.001 |  |  |  |
|  | Poor | Poor | 11 | 20.2 (2.9) | 14.43 (3.4) | <0.001 |  |  |  |
| EDI | Poor | Good | 10 | 83.49 (3.4) | 84.49 (5.3) | 0.137 | 0.184 | 0.057 | 0.312 |
|  | Poor | Mild | 22 | 81.69 (2.5) | 85.03 (0.9) | <0.001 |  |  |  |
|  | Poor | Poor | 11 | 80.96 (3.3) | 82.62 (3.7) | 0.147 |  |  |  |
| EPS | Poor | Good | 10 | 70.8 (6.3) | 69.08 (10.9) | 0.126 | 0.162 | 0.103 | 0.437 |
|  | **Poor** | **Mild** | 22 | 74.33<br>(5.6) | 67.74<br>(1.6) | **<0.001** |  |  |  |
|  | **Poor** | **Poor** | 11 | 75.51<br>(7.6) | 71.86<br>(7.5) | 0.18 |  |  |  |
| **DMC** | **Poor** | **Good** | 10 | 84.22<br>(6.4) | 85.37<br>(9.1) | 0.339 | 0.256 | **<0.001** | **0.004** |
|  | **Poor** | **Mild** | 22 | 78.64<br>(7.2) | 79.02<br>(1.8) | 0.387 |  |  |  |
|  | **Poor** | **Poor** | 11 | 71.66<br>(6.9) | 72.75<br>(6.5) | 0.447 |  |  |  |

**Table 6.** Comparative analysis of gait and EMG indices among participants with mild gait quality at baseline (E1) across responders according to their post-operation gait quality.

| Variable | Gait quality condition before surgery (E1) | Gait quality condition after surgery (E2) | No. | Mean (SD) in E1 | Mean (SD) in E2 | P-Value between E1-E2 | P-Value between Mild_to_Good and Mild_to_Mild in E1 | P-Value between Mild_to_Good and Mild_to_Poor in E1 | P-Value between Mild_to_Mild and Mild_to_Poor in E1 |
| --- | --- | --- | --- | --- | --- | --- | --- | --- | --- |
| **GDI** | **Mild** | **Good** | 15 | 55.99<br>(3.7) | 86.55 (4.9) | **<0.001** | 0.09 | 0.374 | 0.128 |
|  | **Mild** | **Mild** | 21 | 54.41<br>(3.1) | 73.05 (3.4) | **<0.001** |  |  |  |
|  | **Mild** | **Poor** | 21 | 55.39<br>(2.4) | 58.9 (7.2) | **<0.001** |  |  |  |
| **GPS** | **Mild** | **Good** | 15 | 14.56<br>(1.3) | 6.9 (0.8) | **<0.001** | 0.065 | 0.379 | 0.149 |
|  | **Mild** | **Mild** | 21 | 15.19<br>(1.2) | 9.49 (0.8) | **<0.001** |  |  |  |
|  | **Mild** | **Poor** | 21 | 14.77<br>(0.9) | 13.72 (3.2) | **<0.001** |  |  |  |
| **EDI** | **Mild** | **Good** | 15 | 83.41<br>(4.4) | 84.55 (4.9) | 0.291 | 0.138 | 0.14 | 0.448 |
|  | **Mild** | **Mild** | 21 | 81.61<br>(4.7) | 83.21 (5.8) | 0.202 |  |  |  |
|  | **Mild** | **Poor** | 21 | 81.99<br>(4.0) | 83.42 (4.5) | 0.194 |  |  |  |
| **EPS** | **Mild** | **Good** | 15 | 71.35<br>(8.9) | 69.64<br>(10.2) | 0.294 | 0.183 | 0.207 | 0.479 |
|  | **Mild** | **Mild** | 21 | 74.82<br>(10.3) | 71.62 (12) | 0.172 |  |  |  |
|  | **Mild** | **Poor** | 21 | 73.78<br>(8.8) | 70.85 (8.9) | 0.191 |  |  |  |
| **DMC** | **Mild** | **Good** | 15 | 81.91<br>(7.3) | 79.98 (7.5) | 0.252 | 0.112 | 0.293 | 0.225 |
|  | **Mild** | **Mild** | 21 | 78.14<br>(10.3) | 78.32 (8.2) | 0.465 |  |  |  |
|  | **Mild** | **Poor** | 21 | 80.07<br>(8.5) | 80.5 (7.1) | 0.445 |  |  |  |

For patients with severe pre-operative gait conditions (Table 5), all three responder groups experienced significant GDI improvements: 44, 30, and 14 units for good, mild, and poor responses, respectively. While gait and EMG indices (EDI and EPS) at E1 were consistent across these responders, DMC was significantly lower for patients with a poor response (Poor_to_Poor) compared to those with a good response (Poor_to_Good, 72.75 vs. 85.37, p-value < 0.001) and a mild response (Poor_to_Mild, 72.75 vs. 79.02, p-value = 0.004). EDI and EPS also improved from E1 to E2 across all responders, particularly for the mild ones (Poor_to_Mild, p-value < 0.001).

Additionally, the improvement in gait for patients with moderate pre-operative gait issues was significant (Table 6). The mean DMC for the best responders (Mild_to_Good) was higher than that of the other two groups at E1, and the levels of EDI and EPS approached a TD level, although statistical significance was not demonstrated.

Lastly, for patients with the highest GDI levels at E1, while the changes in gait were significant, seven patients (Good_to_Poor) experienced a reduction in GDI (increase in GPS), indicating an unsuccessful surgical outcome. They exhibited a decrease in EDI from 83.73 at E1 to 82.11 at E2, with their DMC being significantly lower than that of Good_to_Good at E1 (78.08 vs. 82.68, p-value = 0.041). Good responders also had a higher initial level of DMC compared to mild responders (78.95 vs. 82.68, p-value = 0.038). Further, the changes in EMG for good responders from E1 to E2 were significant, as measured by EDI (p-value = 0.044) and EPS (p-value = 0.035).

Table 8 also shows the details of the main distal and proximal surgeries for the responders in this study. Dividing the total number of surgeries by the total number of patients for each condition, the average number of surgeries each patient underwent was calculated. To have clinical meaning, we have rounded this number. The better responders with a poor gait (Poor_to_Good) underwent more surgeries (on average 4) than the others (on average 3 and 2). In general, the amount of proximal surgeries was relatively higher than distal surgeries for the patients with a better post-operative gait (21 vs. 15 for Poor_to_Good and 25 vs. 23 for Mild_to_Good).

**Table 7.** Comparative analysis of gait and EMG indices among participants with good gait quality at baseline (E1) across responders according to their post-operation gait quality.

| Variable | Gait quality condition before surgery (E1) | Gait quality condition after surgery (E2) | Number | Mean (SD) in E1 | Mean (SD) in E2 | P-Value between E1-E2 | P-Value between Good_to_Good and Good_to_Mild in E1 | P-Value between Good_to_Good and Good_to_Poor in E1 | P-Value between Good_to_Mild and Good_to_Poor in E1 |
| --- | --- | --- | --- | --- | --- | --- | --- | --- | --- |
| GDI | Good | Good | 30 | 72.06 (7.1) | 86.53 (7.8) | **<0.001** | 0.132 | 0.345 | 0.412 |
|  | Good | Mild | 30 | 70.08 (7) | 71.55 (3.1) | **0.023** |  |  |  |
|  | Good | Poor | 7 | 70.62 (6.7) | 59.72 (3.4) | **0.002** |  |  |  |
| GPS | Good | Good | 30 | 9.85 (1.7) | 6.94 (1.1) | **<0.001** | 0.15 | 0.304 | 0.488 |
|  | Good | Mild | 30 | 10.31 (1.7) | 9.83 (0.8) | **0.021** |  |  |  |
|  | Good | Poor | 7 | 10.29 (1.6) | 13.35 (1) | **0.002** |  |  |  |
| EDI | Good | Good | 30 | 82.45 (6.6) | 85.01 (5) | **0.044** | 0.381 | 0.231 | 0.394 |
|  | Good | Mild | 30 | 82 (5.4) | 82.78 (5.2) | 0.343 |  |  |  |
|  | Good | Poor | 7 | 83.73 (3.5) | 82.11 (3.7) | 0.311 |  |  |  |
| EPS | Good | Good | 30 | 73.71 (13.5) | 68.13 (9.5) | **0.035** | 0.36 | 0.245 | 0.319 |
|  | Good | Mild | 30 | 74.25 (12.7) | 72.19 (11.1) | 0.278 |  |  |  |
|  | Good | Poor | 7 | 70.25 (7.1) | 73.36 (7.3) | 0.356 |  |  |  |
| DMC | Good | Good | 30 | 82.68 (6.9) | 80.91 (7.7) | 0.141 | **0.038** | **0.041** | 0.495 |
|  | Good | Mild | 30 | 78.95 (9.7) | 79.8 (8.6) | 0.37 |  |  |  |
|  | Good | Poor | 7 | 78.08 (4) | 76.28 (8.1) | 0.268 |  |  |  |

**Table 8.** Comparison between the type and number of surgeries for each responding condition.

| Responders | Total number of patients | Distal surgeries |  |  |  | Proximal surgeries |  |  | Average number of surgeries on one patient | total number of distal surgeries | total number of proximal surgeries |
| --- | --- | --- | --- | --- | --- | --- | --- | --- | --- | --- | --- |
|  |  | Achilles tendon lengthening | Baumann procedure | Strayer procedure | bony foot procedures | femoral derotation | rectus transfer | hamstring lengthening |  |  |  |
| Poor_to_Good | 10 | 1 | 6 | 5 | 3 | 10 | 7 | 4 | 4 | 15 | 21 |
| Poor_to_Mild | 22 | 6 | 10 | 9 | 3 | 21 | 11 | 14 | 3 | 28 | 46 |
| Poor_to_Poor | 11 | 0 | 3 | 7 | 4 | 9 | 4 | 1 | 3 | 14 | 14 |
| Mild_to_Good | 15 | 3 | 8 | 8 | 4 | 13 | 6 | 6 | 3 | 23 | 25 |
| Mild_to_Mild | 21 | 2 | 12 | 9 | 9 | 20 | 7 | 9 | 3 | 32 | 36 |
| Mild_to_Poor | 21 | 6 | 7 | 12 | 11 | 17 | 3 | 6 | 3 | 36 | 26 |
| Good_to_Good | 30 | 5 | 16 | 11 | 11 | 20 | 5 | 3 | 2 | 43 | 28 |
| Good_to_Mild | 30 | 5 | 10 | 14 | 16 | 16 | 5 | 3 | 2 | 45 | 24 |
| Good_to_Poor | 7 | 1 | 4 | 2 | 3 | 4 | 2 | 1 | 2 | 10 | 7 |

## 4. Discussion

This study confirms the systematic changes in EMG observed in patients with cerebral palsy following orthopedic surgery. In a relatively larger and more homogeneous cohort, we observed the same findings as Patikas et al. [2] regarding the improvement in post-operative norm-distance for kinetics, kinematics, and EMG (Tables 2 and 3).

The post-operative reduction in soleus activity during initial contact and terminal swing (Figure 2.A) may be attributed to the decreased equinus following the (gastrocnemius) lengthening procedures. This adjustment, common in our population, allowed for a more dorsiflexed position during initial contact. Additionally, the increased activation of the soleus during terminal stance can enhance power generation in the plantar flexors, which is crucial for the body’s forward progression [24].

Moreover, while increased mid-swing EMG activity of the rectus femoris is typically observed in individuals with cerebral palsy [25], the reduction in this activity post-surgery indicates a positive effect on their gait (Figure 2.B). The increase in rectus femoris activity during early stance can also provide sufficient moment to extend the knee in mid-stance. During gait, as the knee extends and the ankle dorsiflexes, the knee moment transitions from an extensor to a flexor moment, allowing the quadriceps to cease contracting and the ankle to absorb power through the eccentric contraction of the gastrocnemius–soleus complex [26]. These synergistic changes observed in our study in the EMG of the soleus and rectus muscles may result in an improved plantar flexion–knee extension coupling mechanism [26], leading to better gait quality as indicated by the GDI and GPS (Table 2).

The examined EMG indices, EDI and EPS, basically describe EMG patterns, and tend to become more typical (Table 3) as gait becomes more typical after the surgery (gait parameters, Table 2). This might be interpreted as a biomechanical aspect of EMG activity. In contrast, DMC seems to be a quantity that is relatively independent of the changes in gait pattern or indexes derived from them (Table 3). Therefore, and this is the second hypothesis we examined in this study, DMC can be a partial predictor of outcomes: ’Good DMC = good response; Poor DMC = poor response’ (i.e., of orthopedic/biomechanical intervention). According to [18], we applied a clustering algorithm to divide the subjects into groups of patients with similar gait quality in E1 and the same response to the treatment in E2.

Considering the possible recovery conditions, DMC was the only measure that showed a difference between the responding groups with the same baseline GDI (Tables 5, 6, and 7), while it remained the same from E1 to E2. The patients with a relatively low DMC (≈ 70 out of 100 for TD) along with poor gait quality (GDI ≈ 40 out of 100 for TD) are more likely to have worse outcomes after treatment (Table 5). This finding interestingly implies that orthopedic surgeons should be cautious not to overtreat patients with severely limited motor control. Conversely, a relatively high DMC (≈ 80 out of 100 for TD) for patients with a better pre-treatment GDI (≈ 70 out of 100 for TD) can lead to further improvement in their gait following the intervention (Table 7). Moreover, for mild cases (Table 6), it can also be seen that the average DMC for the good responders was higher than for the other groups. However, for these cases, it might be difficult to distinguish between the biomechanics and the motor effect on their gait deficit. It is crucial to acknowledge that the descriptive terms used in this study, such as poor, good, mild, better, and worse, are context-specific and pertain to the population included in our research. In other clinical settings, where the severity levels of patients may differ, these thresholds might not apply uniformly. Nevertheless, the inclusion of 167 patients in our study provides a sufficiently large sample size to generalize the systematic changes in EMG post-surgery, validate the application of EMG indices in clinical practice, and underscore the significance of DMC as a measure of motor control function in clinical decision-making. While prior studies addressed pre-treatment femoral anteversion [27], knee flexion [28], dynamic hip flexion [29], and gait profile score [30] as predictors of post-operative outcomes, this is the first study to examine the connection between DMC and gait following surgery in a relatively homogeneous group of patients with CP.

Comparing the type and number of surgeries among the responders, it appears that those with poor pre-treatment gait quality who demonstrated better responses underwent the most extensive surgical interventions. Furthermore, proximal surgeries, such as rectus transfer, hamstring lengthening, and femoral derotation, seem to have resulted in higher responder rates compared to distal surgeries, such as Baumann-Strayer and bony foot procedures, for patients with initially poor and mild gait quality. This may introduce a potential bias in the grouping methodology employed in this study. Our research primarily focused on the applicability of EMG as a clinical measure to enhance decision-making for patients with CP. However, we recommend that future researchers conduct more focused studies on the impact of different surgical approaches on EMG changes and their relationship with gait improvement. Additionally, the influence of growth, changes in muscle mass, and spasticity over time, particularly following surgery, on the EMG and gait of patients with cerebral palsy, should also be explored in future studies.

## Data Availability

All relevant data are within the manuscript and its Supporting Information files.

## 6. Author Contributions

MD: Writing original draft, Data analysis; FS: Review & editing, Methodology; RR: Data analysis; DAP, NAB and KG: Review & editing; SIW: Review & editing, Conceptualization, Methodology, Project administration.

## 7. Data Availability

The data supporting the conclusions of this article will be made available by the authors upon request.

